# Domestic abuse, primary care and child mental health services: A systems analysis of service coordination from professionals’ perspectives

**DOI:** 10.1101/2024.10.15.24315525

**Authors:** Claire Powell, Olumide Adisa, Lauren Herlitz, Shivi Bains, Sigrún Eyrúnardóttir Clark, Jessica Deighton, Shabeer Syed, Ruth Gilbert, Gene Feder, Emma Howarth

## Abstract

**Objective:** We explored how services work together to support parents and children experiencing both parental intimate partner violence (IPV) and parental or child mental health problems by drawing on the perspectives of professionals working in primary care, children and young people’s mental health services (CYPMHS), and domestic abuse services.

**Methods:** We conducted a qualitative study, interviewing 38 professionals in three geographically contrasting local authority areas in England. We carried out framework analysis using a systems approach and mapping techniques to understand the service interrelationships and boundary judgements of professionals.

**Results:** The relationships between domestic abuse services, CYPMHS, and primary care were complex, involving funders and commissioners, local authority strategic groups, and wider services such as schools and children’s centres. Participants consistently identified a gap in the relationship between statutory CYPMHS and domestic abuse services. Other service gaps were for children living with ongoing or intermittent IPV and for children and parents with needs falling below or between service thresholds. There was a gap in support services for users of abusive behaviour to prevent future IPV. An overview of staff perspectives revealed differing views on treating the effects of trauma, and the co-ordination and sequencing of care.

**Conclusion:** Improving the response to children and adults experiencing mental health problems in the wake of IPV requires a systems perspective to understand the barriers to service co-ordination. Our findings indicate a particular need to address the gap between CYPMHS and domestic abuse services. Current ways of working with adults could be adapted for children, in addition to learning from examples of best practice in the study sites.

## Introduction

Globally over one in four women are reported to have experienced intimate partner violence (IPV) in their lifetime (1) and in the UK it is estimated that one in five children have been exposed to parental IPV. (2) Intimate partner violence refers to physical, sexual or psychological harm, including threatening, aggressive and coercive behaviours, by an intimate partner or ex-partner. (3) In the UK, children living with parental IPV are now acknowledged as primary victims of abuse. (4)

The impact of IPV on parental and child mental health is well documented, (5–7) as is the associated increased use of healthcare services by survivors of IPV. (8) Findings from the UK’s ALSPAC longitudinal study show that parents and their children who had experienced IPV were more likely to experience depression than families that were not exposed. Children who experienced both parental IPV and maternal depression were themselves more likely to experience depression than their peers who were exposed to only one of these parental factors. (9) Similarly, parents with IPV recorded in English electronic healthcare records in the first 1000 days of their child’s life were more likely to have recorded mental and physical health problems than parents with no recorded IPV. (10)

In recognition of the links between family health outcomes and parental IPV, UK policy emphasises the importance of partnership working between health services and domestic abuse agencies, along with clear referral pathways for victims of violence and abuse and a co-ordinated local responses. (11) National Institute for Health and Care Excellence (NICE) guidance suggests a multi-agency approach to IPV (12) but in general, current policy and guidance is focused on the response to adult victims with more limited advice as to how this should be operationalised for working with children.

Although multiagency approaches for parental IPV are advocated, evidence from practice shows siloed provision for adult and CYP mental health and IPV. (13) Reported factors contributing to this include: challenges in multi-agency co-operation for children with child protection concerns (including parental IPV); (14) healthcare infrastructure; (15) and administrative issues affecting referral pathways, funding and multi-agency miscommunication. (15,16)

A gap in specialist support for children exposed to IPV is well recognised. (17) Research into outcomes for children living with IPV found that most interventions for children are delivered by the voluntary (or non-profit) sector. (16) Even where IPV services, such as IRIS, (18) are linked to UK general practice (family medicine) there are no recommended approaches for how either physical or mental health services should identify children’s needs or co-ordinate support when children also have mental health problems.

Previous research has criticised health services responding to IPV for taking a narrow, single sector perspective, suggesting a systems approach would be useful. However, recent research identifying systems factors that increase screening and identification of IPV in healthcare settings did not include children. (19) There is also a lack of research on the ‘enabling conditions’ (20) or systems barriers to service co-ordination specifically for families experiencing mental health problems and IPV.

We addressed the following research questions using a systems approach: 1) How do domestic abuse, primary care and child mental health services co-ordinate services for families experiencing mental health problems and IPV? 2) What are professional perspectives of how services should support families experiencing mental health problems and IPV? 3) How do professionals perceive service boundaries for families experiencing mental health problems and IPV?

## Methods

We used a qualitative study design to understand how primary health care, child mental health, and domestic abuse services work together to identify and support families experiencing parental IPV and mental health problems (in either parent or child or both). We followed the Consolidated Criteria for Reporting Qualitative Research (COREQ) (See supplement 1) and we pre-registered our protocol. ([OSF Registries | Intimate partner violence and mental health of children and parents or carers: qualitative study protocol](https://osf.io/4ynrh)) Changes to the protocol are detailed in supplement 2 and see supplement 3 for our glossary and definitions.

We carried out 36 semi-structured interviews between November 2022 and February 2023 with 38 professionals in three areas of England. Participants were asked to describe a family’s journey through their services, including how mental health problems and IPV were identified and recorded, support provided, and coordination of inter-service relationships.

### Public & Patient Involvement

We consulted our lived experience advisory group at the study design and data analysis stages. The group contributed questions to the interview schedule and highlighted areas of importance in the analysis. In addition we formed three professional advisory groups, one for each recruitment area. These groups input into the interview schedule design, supported with participant recruitment, and fed back on the findings.

### Theoretical framework: systems approach

We used Meadows’ definition of a system for this study: a system typically comprises of “three kinds of things: elements, interconnections, and a function or purpose” and “is more than the sum of its parts” (21). Therefore, a system can be characterised or perceived as a set of relationships in which the elements when taken together form a whole. The whole itself may be part of wider systems with their own set of elements that are linked to other systems. As our focus was on service co-ordination, we were particularly interested in the interconnections between elements (i.e. relevant health and social care services).

We also drew on Ulrich’s’s theory of boundary critique which suggests that it is important to access a ‘diverse variety of stakeholder views in defining problems and complex boundaries’. (22) In our study, we were interested in service boundaries, i.e. who was perceived as eligible for and able to access services.

### Ethics

We did not ask participants about personal experiences of IPV; however we were aware the subject matter may have been difficult or that participants may themselves have experienced IPV. Participants were able to feedback on the interview process and they were offered a debrief sheet with contact numbers for support services. Our study was approved by UCL Research Ethics Committee (17893/003) and NHS Health Research Authority (315188).

### Sampling & Recruitment

The study areas were based on pre-existing contacts from an earlier study conducted in England. (42) We aimed for contrasting areas based on geography but also willingness from a key contact to provide support. Our final areas were 1) a northern, primarily rural local authority; 2) a western, city council local authority; 3) an inner-city London local authority.

In consultation with the study advisory groups we designed a maximum variation sampling framework to recruit participants from a range of professional roles (including frontline, managerial, and strategic/commissioning) across primary care (n=6), child mental health services (including voluntary sector and statutory sector covering universal to specialist services) (n=13), and domestic abuse services (n=19) in each area. We used the framework to target specific professionals, but we included other professionals if they expressed an interest and met our criteria. See Table 1 in below for participant demographics. (See also Table 2 in supplement 4 for sampling framework, recruitment targets and final numbers, and supplement 3 for definitions of professional groups.)

**Table 1.**
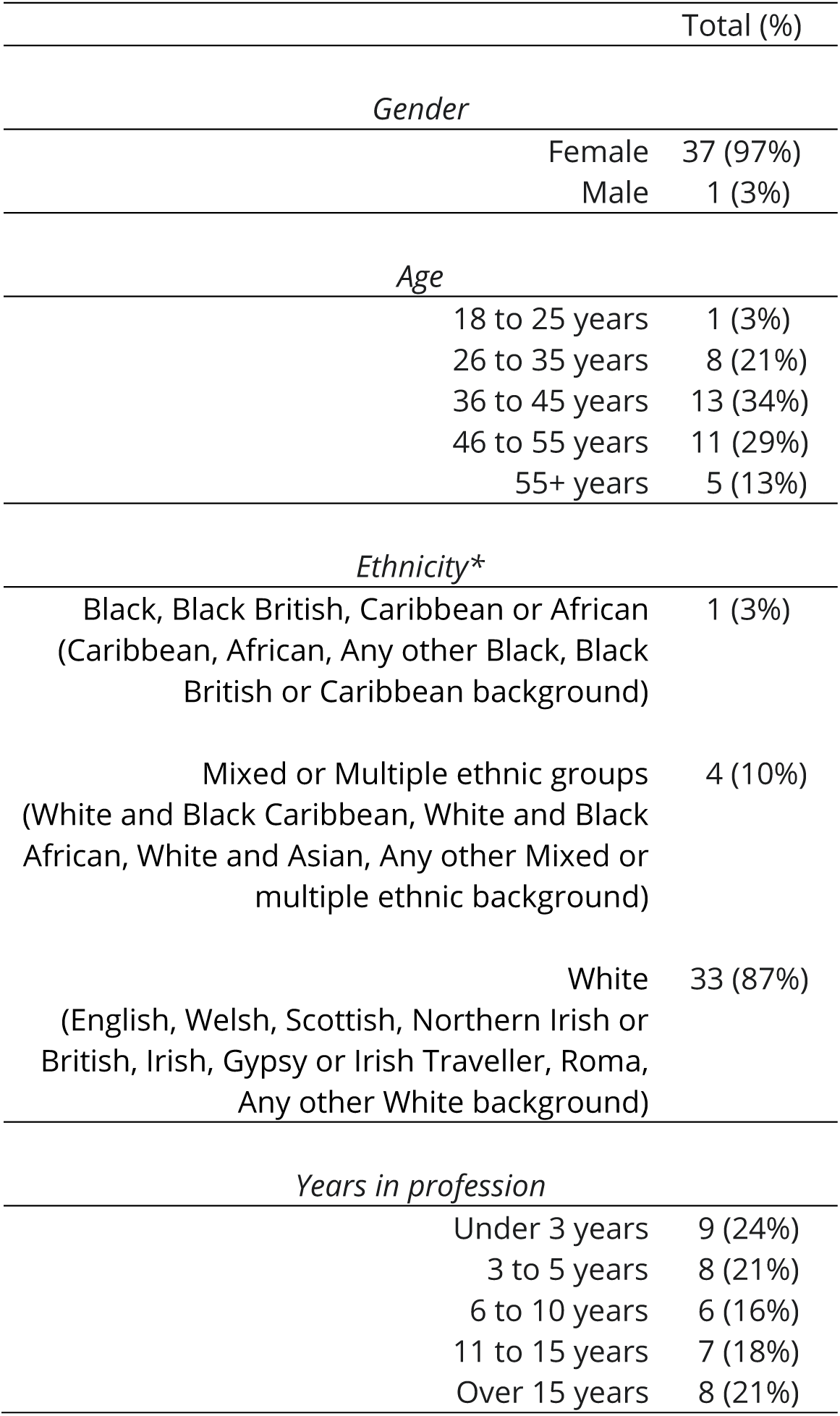
Interview participant demographics.

### Procedure

Advisory group members emailed the study information to relevant organisations or individuals and interested participants contacted the study team directly. We also used snowball sampling, asking interview participants to suggest suitable colleagues. Once a potential participant made contact, we shared the information sheet, written informed consent form and brief demographics questionnaire in REDCap, (23) and organised an online interview in Teams. 43 professionals contacted the research team and 7 (16%) did not take part in an interview.

We offered individual 60-minute interviews (n=31) and 90-minute group interviews (n=2) where there were two or more participants from the same organisation. For GPs, we offered a shorter 30-minute individual interview (n=3) to encourage participation. We adapted the interview schedule for each type of role and all participants were sent the interview questions in advance.

Interviews were carried out by experienced female qualitative researchers (SC, LH, CP) who met weekly throughout the interview period to discuss any concerns and track recruitment progress. Interviews took place online and were audio-recorded in Teams and sent securely to a professional transcriber for transcription. Participants were offered a shopping voucher for their time and the opportunity to review summary findings.

### Analysis

While data were being collected, each interview was summarised in an area-specific rapid assessment procedure (RAP) sheet (24) (a structured template) to facilitate initial analysis. This enabled real-time consideration of possible themes, as well as rapid cross-case comparison, and reflection on new probes or follow-up questions. The RAP sheets were used to create area-specific summaries, which were sent to all participants and advisory groups for optional feedback after data collection was completed.

Following the feedback and team discussion we carried out framework analysis. (25) After the initial inductive indexing and coding of the data in NVivo 12 by one researcher (CP) (26) we applied our systems lens to organise the codes and structure the analysis of themes. We used the codes to create system maps in Kumu (27) to visualise our interpretations related to organisational relationships. Initial stages of analysis were discussed with the interview team, the wider research team, and the lived experience group to refine the themes and the analysis.

## Results

We explored how primary care, domestic abuse, and child mental health services in England work together and we developed three broad themes: 1) the interrelationships of organisations and cross-sector groups involved in care for children and parents experiencing mental health problems (MHP) and IPV; 2) the boundaries of care between services; and 3) tensions in perspectives on how CYP/parents should be supported.

### 1. INTERRELATIONSHIPS OF ORGANISATIONS AND CROSS-SECTOR GROUPS INVOLVED IN CARE FOR CHILDREN AND PARENTS EXPERIENCING MHP AND IPV

We identified three groups of organisational interrelationships that affect how primary care, domestic abuse and child mental health services work together to support families: A – funding-commissioning relationships; B – interface relationships (i.e. between frontline service delivery and funding/commissioning); C – inter-service relationships. For each group we present a systems map based on the interview codes and thematic summaries.

#### A Funding and commissioning relationships

Participants highlighted the key role that funding and commissioning play in service co-ordination for families. The three core aspects of funding and commissioning relationships were: national government budget allocation; wider commissioning relationships which support or hinder service delivery; individual commissioners and what they do. See Figure 1 for a representation of these relationships.

**Figure 1.**
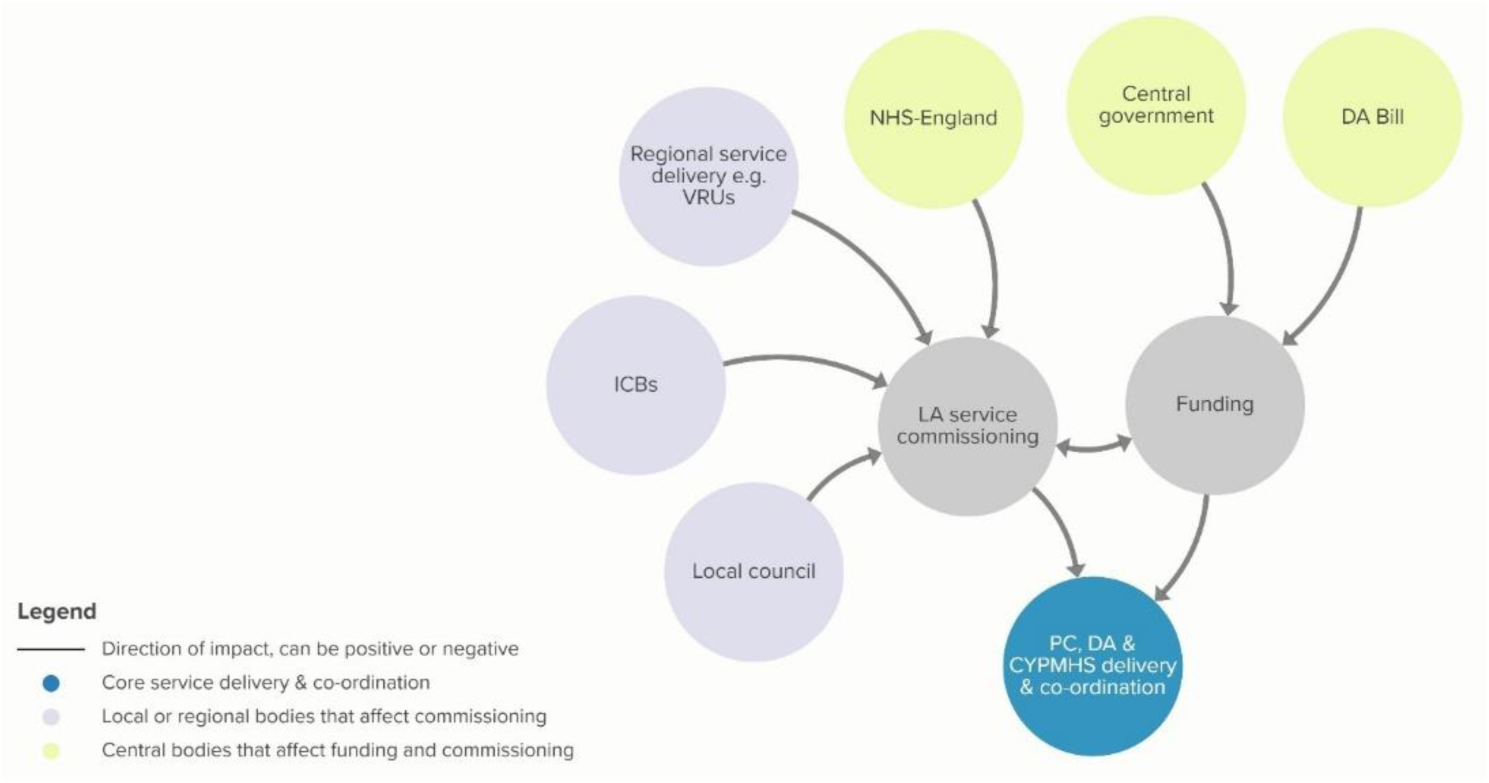
Map of funding and commissioning relationships

##### A.1 National government budget allocation

The Domestic Abuse Act (2021) was cited as a positive piece of legislation which has made provision for specific funding of children’s domestic abuse support services; this was seen as potentially funding some otherwise unavailable mental health support for children and increasing domestic abuse services’ capacity to work with other services. However, service commissioners and leaders pointed out that the use of national government funding is often restricted and does not always allow sufficient time or flexibility to create the service co-ordination needed, for example, to implement joint commissioning and contracting. Furthermore, we repeatedly heard from professionals that the current decade was one of the worst periods of funding for public services they had experienced in their careers, and this was limiting and reducing available services that children and families could access and their capacity for working together:

> “All of us, schools, CAMHS, social care, [are] all very underfunded, very fractured kind of services and we’re all trying to come together and … it’s like a broken piece of pottery and you’re trying to put all the pieces back together and all the bits aren’t really fitting” (P10, CYPMHS worker)

Other participants discussed this fragmentation and explained that lack of funding meant that only less intensive wellbeing work, often in groups, could be funded, not the in-depth one-to-one mental health support that parents and children needed.

##### A.2 Wider commissioning relationships

Participants discussed a variety of commissioning relationships which might support or hinder service commissioning and co-ordination. Joint commissioning arrangements with bodies such as NHS England and Integrated Care Boards (ICBs) were seen as supportive and productive, crossing the health-social care interface, and enabling relationships between services. When local councils provided support for domestic abuse as a political priority, this enabled commissioners and service leaders to create innovative services. However participants explained that regional bodies, such as Violence Reduction Units (VRUs), might be separately commissioned to deliver IPV services funded by central government. When these bodies did not communicate with local commissioners or delivered their programmes in parallel, this negatively affected local service co-ordination resulting in service gaps or duplication:

> “It’s not just about receiving the money; it’s about how we’re managing those resources. You know, implement something that’s a little bit of a bigger scale, that’s got economy of scale, that will provide the same services across bigger patches. But this is the problem, when we see things being delivered pan-[region], … commissioners, locally, have got very little say around it and that’s the problem.” (P13, Domestic Abuse)

##### A.3 Individual commissioners and what they do

Local authority service commissioners’ prioritisation (or not) of domestic abuse, their knowledge and expertise, cross-sector networks, and capacity were considered by participants to directly affect inter-service co-ordination:

> “Maybe commissioners who’ve got 70 different portfolios and DAs just a little drop in the ocean, would they have the time and capacity, would they prioritise?” (P13, Domestic Abuse)

According to our participants, commissioning effective services that worked well together depended on vision, commissioning relationships across sectors, and the integration of administrative procedures such as joint contracting and shared case management systems. Without these things in place, mental health problems in the context of domestic abuse could be overlooked because it did not seem to be anyone’s specific responsibility.

#### B Interface relationships

There were three types of relationships at the interface of service delivery and funding-commissioning that participants described where primary care, child mental health and domestic abuse services might cross paths and could help or hinder cross-service working. These were: 1) local area strategic groups; 2) safeguarding partnerships; 3) domestic homicide reviews. These were all affected by funding-commissioning relationships and directly shaped service delivery relationships. See Figure 2 for a map of these relationships.

**Figure 2.**
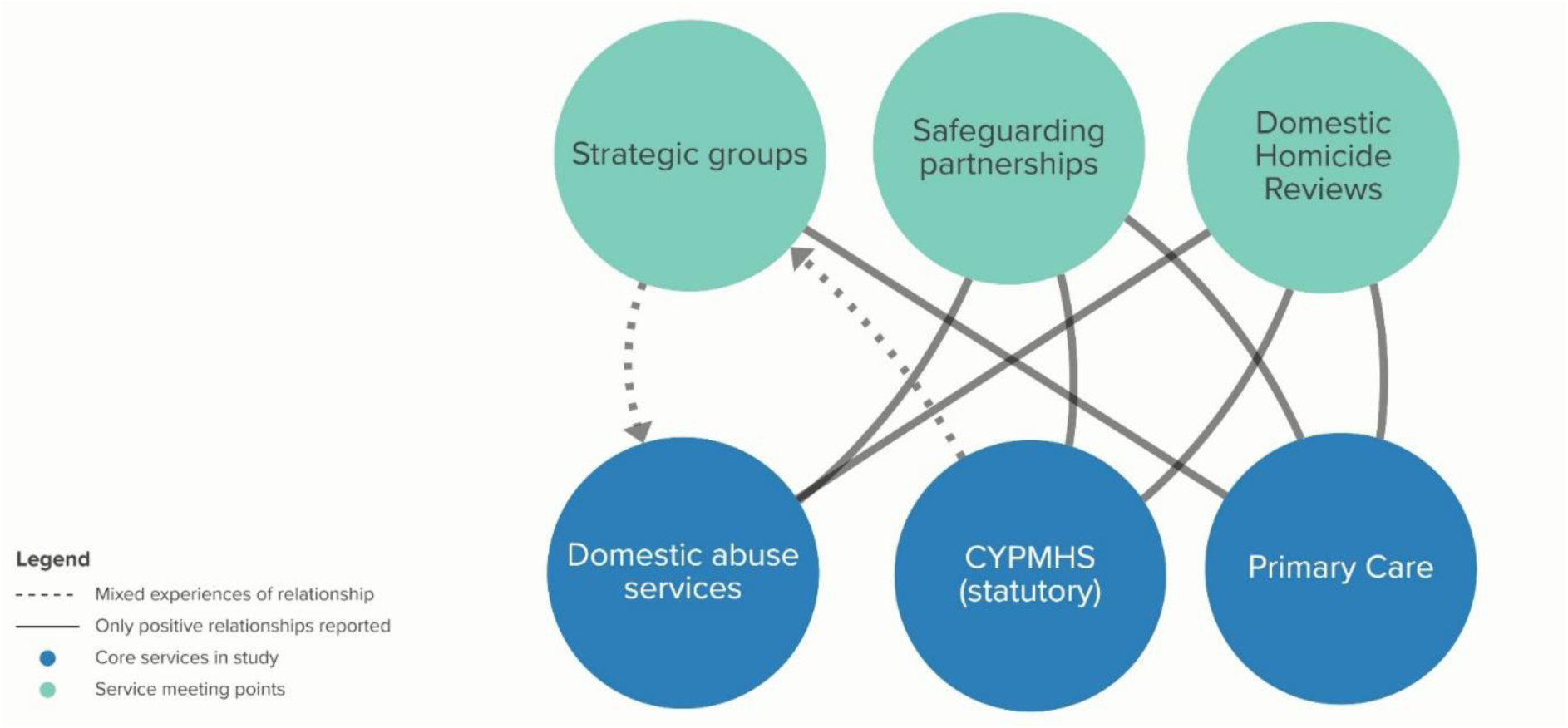
Map of interface relationships

##### B.1 Local area strategic groups

Strategic groups (including both steering and operational groups) bring together different partners and can facilitate joined-up support pathways. These groups were described as part of the commissioning process to oversee service delivery and partnership working and were seen as key to co-ordinating support between services. However, member attendance and inclusion were key to their effectiveness, as reported by interviewees. A common complaint was the non-attendance of mental health services (despite being invited):

> “I’d say one of the biggest gaps in all these meetings I’ve been talking about is representation from [mental health trust]. … And certainly CAMHS… I’ve never seen them strategically round the table, at all.” (P14, Domestic Abuse)

In addition to this we heard about some domestic abuse services and relevant (particularly small) charity sector service providers not being invited:

> “At the minute we don’t have a voice there [Domestic Abuse Board] …if you think about the 400 plus people we worked with last year, their voices are not being represented. I’m really keen for us to have a seat, so that we can have those conversations with other services, we haven’t had strong links with health, so that’s an area that we really want to develop and make some links there.” (P15, Domestic Abuse)

The exclusion of domestic abuse services, including specialist ‘by and for’ services was also highlighted in discussions around multi-agency meetings. Participants in strategic roles across health and domestic abuse suggested this might add to the communication challenges between voluntary sector domestic abuse (and related) services and statutory CYPMHS.

##### B.2 Safeguarding partnerships

Safeguarding partnerships were seen to help joined up working and as an opportunity to think about the whole family, as well as encouraging multi-disciplinary working. It was a key structure for bringing awareness of domestic abuse into health services and for multi-agency referrals. Where there was limited cross-working between domestic abuse, primary care and child mental health services, safeguarding was identified by some interviewees as the only arena where these services might work together because of their clear purpose:

> “It’s within safeguarding we contribute to how those services work together… safeguarding should be a thread through all those services and how they work together.” (P16, Primary Care)

However some small charity sector organisations, especially by and for services, felt they were not always seen as equal partners in the process:

> “The only thing I would say is we’re not always or experiences have been at times where we don’t get the notes, so we won’t get the sort of notes or minutes like others will, we’re just sort of called in and … not necessarily, I suppose, seen as equals in relation to other.” (P24, Domestic Abuse)

##### B.3 Domestic Homicide Reviews (DHRs)

DHRs were also seen as an opportunity to improve service co-ordination, often highlighting poor service communication or challenges identifying families at risk. Some participants from the domestic abuse sector highlighted that they had only been able to develop positive relationships with mental health services after a DHR had flagged issues in cross-service communication.

#### C Inter-service relationships

Participants reported that referrals were the primary relationship between services. In this section we describe the nature of relationships between domestic abuse, child mental health and primary care services, as well as the impact of wider services. Figure 3 shows the wide eco-system of services in which families experiencing both domestic abuse and mental health might access support.

**Figure 3.**
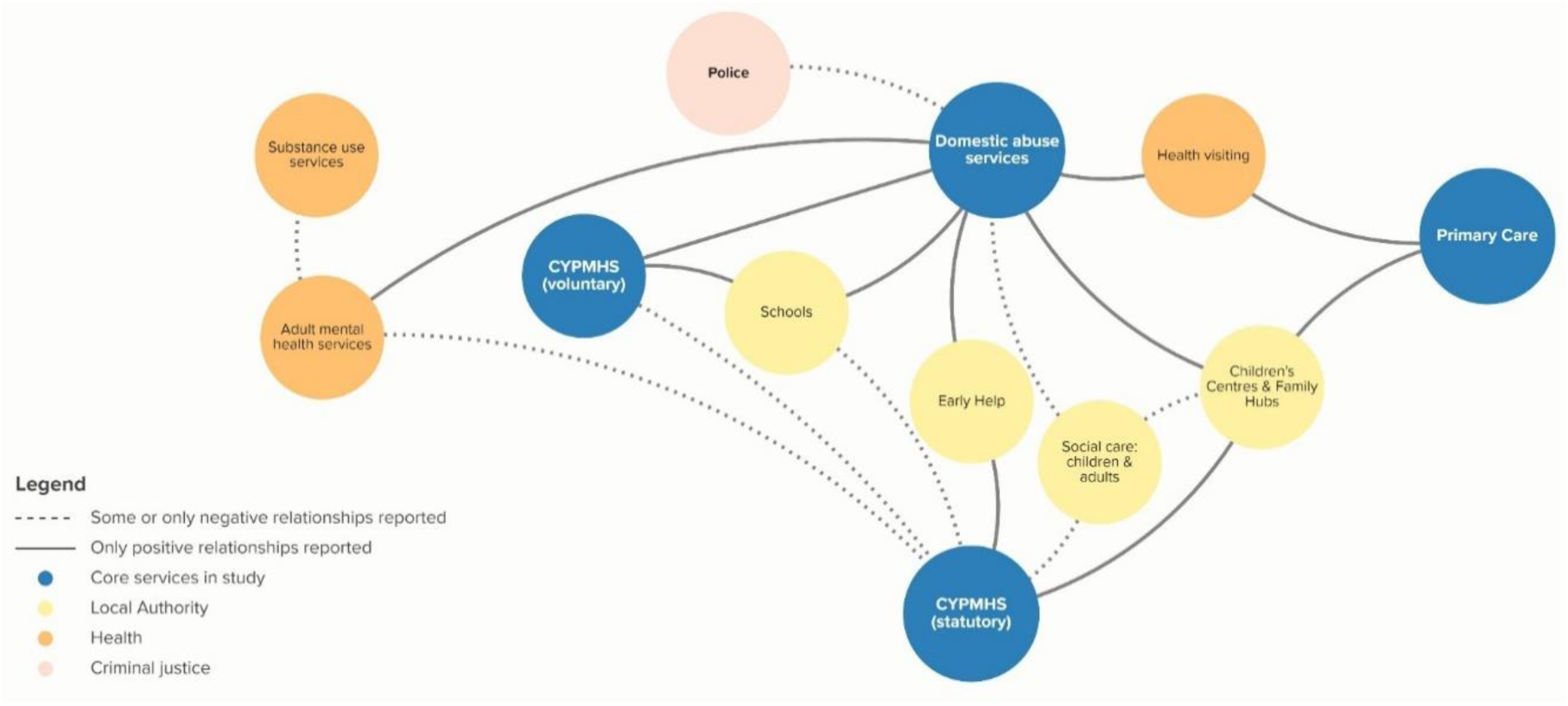
Map of inter-service relationships

##### C.1 Connecting services between domestic abuse, CYPMHS and primary care services

According to our interviewees, children’s centres and family hubs seemed particularly well-integrated with domestic abuse, CYPMHS and primary care services. Early help services (for families who don’t meet the social care threshold but need additional support) appeared to have good connections to both domestic abuse services and CYPMHS. Small voluntary sector organisations reported that they needed visibility from statutory services (i.e. advertising and referrals) to be well-integrated in the local service offer.

The main gap that professionals described (and found challenging) was that between domestic abuse services and statutory CYPMHS, perhaps related to the challenges identified in funding, commissioning and interface relationships described above. One participant described it as a ‘wall’ (P17, domestic abuse), another as ‘very hit and miss with a lot of misses’ (P24, domestic abuse), and in general the relationship was described as mediated by social care or schools. Participants across domestic abuse services reported that in their areas they were unable to refer to mental health services directly without the support of another local authority service (e.g. a school) or a healthcare professional.

Primary care services seemed less well-integrated with wider local authority services. However, GPs described positive relationships with health visitors which enabled them to be alerted to families with children under 5 years exposed to parental IPV. There did not seem to be a similar relationship with community-based professionals for older children. Whilst GPs described making referrals to CYPMHS for young people exposed to IPV, there was rarely any other contact between the two services. One participant described the primary care role as:

> “We are the holder of the information… our important thing is to make sure it’s all visible and it’s reviewed and it’s there for other people to see.” (P18, Primary Care)

Other participants felt there previously had been more opportunity for conversations with GPs around mental health support for young people and parents, but this was no longer possible given increased caseloads in primary care.

##### C.2 Support from the wider network

Participants described the impact of poor service co-ordination in the wider network (i.e. beyond primary care, domestic abuse and CYPMHS) and how this affected support for families with a range of needs. A particular issue was the lack of communication between adult mental health services and substance misuse services and the practice of excluding parents from accessing mental health services until substance misuse was addressed. This could leave parents at greater risk of harm from their partners using abusive behaviours, less likely to be able to address their issues, as well as having a negative impact on their children.

Participants explained how disagreements between social care services and Family Hubs about categorising parental relationships as conflict or domestic abuse might mean families were referred to inappropriate support, leaving families unable to access domestic abuse or CYPMHS services when needed. Different definitions of IPV and inter-parent conflict across services seemed to underpin this, although one participant speculated that categorising parental interactives as conflictual rather than abusive enabled cases to be passed from social care to other services, which might help reduce the high caseloads in social care.

### 2. BOUNDARIES OF CARE BETWEEN SERVICES

Participants discussed how the threshold criteria for access into CYPMHS and adult mental health services affected families experiencing IPV from accessing timely mental health support. Four groups of children and families referred for mental health problems were identified by practitioners as particularly affected by access criteria: children who live with current/ongoing domestic abuse; children below social care safeguarding thresholds; parents/children with mental health presentations that fall between the primary and secondary mental health care thresholds; and mothers with high needs and custody of their children. Practitioners discussed the impact of exclusion from services for families and the implications for service co-ordination.

#### 2.1 Children living with domestic abuse (i.e. still living with the perpetrating parent)

Interviewees perceived that many mental health service criteria exclude children who are not considered to be living in a situation of safety:

> “One of the … exclusion criteria from many of the young people’s mental health service response, is needs stability, they need to be safe, there cannot be ongoing abuse, and of course that excludes every child that is experiencing domestic abuse, living in a refuge, mums in a coercive controlling relationship. They’ve got high level trauma symptoms, they’re of course demonstrating harmful behaviours in school environments, and that is classified as not safe or stable enough to work with generic mental health services.” (P17, Domestic Abuse)

Participants from domestic abuse services and both statutory and voluntary sector child mental health services reiterated this, highlighting the number of children with mental health needs who could not access support for this reason.

#### 2.2. Children below social care safeguarding thresholds

Children perceived as living in an abusive situation that is not risky enough to meet social care thresholds were also considered to be less likely to access CYPMHS even with significant mental health needs. Interviewees acknowledged this was because of reduced funding and service cuts which meant that CYPMHS thresholds are only really met by children also meeting social care thresholds. As one primary care practitioner explained:

> “The threshold for CAMHS is so high that a child really needs to be in quite significant distress, and if they were in that much distress and there was that much, you know, there was that much of a need, I would say nine times out of ten they’re already involved in social care.” (P11 Primary Care)

#### 2.3 Parents/children who fall between the primary and secondary mental health care thresholds

In England, primary mental health care provides access to ‘NHS Talking Therapies’ (previously known as ‘Improving Access to Psychological Therapy’ or ‘IAPT’), which predominantly comprises CBT interventions. Interviewees were concerned that adults and children exposed to domestic abuse are considered too complex for this level of therapy and that standard CBT as opposed to trauma-focused CBT is often inappropriate for trauma survivors. However these same families did not then meet the diagnostic criteria or thresholds for higher tier or specialist services; this left them unable to access any statutory mental health support as described by this practitioner:

> “I would say 50% of my cases, I go back to the GP and say, you know, the NHS talking therapy services isn’t enough for this client, but then they don’t meet the threshold for secondary mental health services, so we’re kind of stuck because I can’t do much work while she’s in crisis, but there are no resources to help her come out of crisis.” (P11, Primary Care)

#### 2.4 Mothers with high needs

The final group refers to mothers with high level needs including mental health problems and substance use problems, who have custody of their children. Whilst there is some specialist mental health and domestic abuse provision for mothers who no longer have custody of their children, mothers with children are unable to access residential domestic abuse provision with high-level mental health support.

#### 2.5 The impact of children and parents being excluded from accessing services

The impact on services and families of these threshold criteria and lack of support for some children and families was described by this practitioner:

> “I think the waits are just so, so severe and the child has to be so disabled, by the time they get seen by CAMHS in, sadly, you just feel that sometimes you think a stitch in time would save nine and I fear that, that there often isn’t that possibility, there isn’t that capacity in the service to see these children before they do get completely unravelled.” (P19, Primary Care)

Several participants explained that children would deteriorate on lengthy waiting lists. Parents were also more likely to return to abusive partners if they were unable to access timely and appropriate support. Wider support services described how they might avoid even making referrals to mental health services because they did not want families to be rejected:

> “What we don’t want is families being bounced around, being told … you’re not relevant for this service… People don’t like to hear that and that has ramifications down their life, to be told that they’re not complex enough or too complex.” (P20, Primary Care)

Voluntary sector services and primary care services described how they tried to fill the gap by keeping families on for longer in their own services, recommending private therapy or trying to find alternative support through schools. The boundaries of care between services seemed both to reflect funding and commissioning arrangements (and their gaps) but also practitioner perspectives on who should be supported and how in different services.

### 3 TENSIONS IN PERSPECTIVES ON HOW FAMILIES SHOULD BE SUPPORTED

On analysing interviewee perspectives of how care should be provided, there were three points of divergence: 1) the problem with ‘safety first’; 2) the role of child and young people mental health services; 3) who is responsible for users of abusive behaviour.

#### 3.1 The problem with ‘safety first’

There was general agreement that mental health interventions for parents and children were limited when environmental factors (such as poor housing) are not addressed, and that trauma responses cannot be processed while traumatic experiences are ongoing. However, participants outside of mental health services were keen to point out that it is difficult to keep people safe if they are mentally unwell or distressed:

> “A really big barrier for most women to make positive decisions about relationships, you know, having the strength to make decisions to leave relationships, … being able to prioritise their children’s needs… really whittles down to their capacity and mental health. If a mum is in complete mental health distress, then it’s really difficult for me to put all these options out … if they’re in crisis, none of those things are really easy to take on board and to action.” (P11, Primary Care)

Domestic abuse services felt they were equipped to offer emotional support, mental health safety planning or advocacy around service access, but not therapy or formal mental health interventions. Domestic abuse services staff – in particular, advocates with a focus on practical support – described how it was difficult not to be pushed into a therapist role when providing support. Staff felt that approaches which brought in consultation and supervision from mental health specialists in domestic abuse services would be useful to negotiate some of these challenges.

#### 3.2 The role of child and young people mental health services

The next diverging perspective was around the role of CYPMHS for families experiencing domestic abuse. Many domestic abuse practitioners felt CYPMHS saw domestic abuse primarily as a safeguarding issue rather than as a risk for or contributor to mental health problems:

> “I guess is it the case that mental health services don’t really understand domestic abuse and sexual violence, and maybe they are only seeing it in terms of safeguarding of their own clients, rather than how could they be working in prevention, how could they be working in crisis, and how could they be working in recovery?” (P12, Domestic Abuse)

This participant felt mental health services could be part of domestic abuse prevention and recovery for children because they are already in contact with so many children who have experienced parental IPV.

There was a perception that there is a mismatch between CYPMHS thresholds and diagnostic criteria, and children’s presentations when affected by trauma (in this case, domestic abuse), i.e. that children could experience severe mental distress and need support, but this did not ‘fit’ service criteria. Child mental health staff from both the statutory and voluntary sectors agreed that short-term interventions were unsuitable for addressing the impact of domestic abuse, and that there were not the resources in the public sector for children impacted by trauma.

However, domestic abuse staff and voluntary sector children’s mental health staff felt that there were some more fundamental differences between themselves and statutory child mental health practitioners’ perspectives on mental health. The conflict seemed to centre on whether trauma-related mental distress is different from ‘mental illness’ – with a perception that domestic abuse services and the voluntary sector had a trauma-informed approach to mental health, whilst mental health services did not.

Some participants suggested that where statutory child mental health services were aware of domestic abuse, their role was providing support for children with historic trauma, “we’re in the game of processing past trauma” (P21, CYPMHS), whilst it was seen as the voluntary sector’s role to provide support with current trauma. In addition there was a general perception that statutory child mental health services are less aware of the complexities of domestic abuse, particularly around coercive control, jealousy, stalking and harassment or challenges facing people trying to leave abusive relationships.

We identified from the interviews that domestic abuse services were systematically asking as part of their intake assessments about the mental health of parents and children, whilst statutory child mental health services were not systematically asking about or identifying family IPV. This was reflected in levels of training staff had received too – with all domestic abuse staff reporting some mental health training, whilst few child mental health practitioners had received IPV training, and notably most of those who had were based in the voluntary sector. (See supplement 5 for further details.)

Participants agreed that there is a lack of clarity about who is responsible for what support at what time for families experiencing domestic abuse – in particular, around trauma support, mental health interventions, counselling, and referral pathways in general. Participants from both sides agreed that they wanted to address the silo between domestic abuse services and CYPMHS. CYPMHS practitioners discussed how difficult it was for them to identify domestic abuse if children did not talk about it and accompanying parents did not provide this information.

#### 3.3 Who is responsible for users of abusive behaviour?

The final diverging perspective was around support for users of abusive behaviour (usually fathers) to change their behaviour. There was general agreement that there is a gap in support for parents who use abusive behaviour and young people who may begin to in their own relationships. Some participants felt their support of children and the non-harming parent was limited if there was no support available for the parent using abusive behaviour to end their abuse. There was a sense that parents who use violence are ignored by services, in particular social services and mental health services, and that outside of specialist perpetrator programmes there is a lack of responsibility for and skills to work with this group of parents.

Some specialists described how colleagues might be frightened of parents who use abusive behaviour or worry about upsetting them (and the impact on the family). However this fear resulted in avoidance of any contact with the parents who use abusive behaviour:

> “I was worried about if I said something that made this man very angry that he would then actually seriously hurt his family or kill them.” (P23, CYPMHS)

Specialist domestic abuse workers wanted mental health services to be able to hold perpetration and mental health in mind simultaneously, both for the parent who uses violence and for young people who start to display abusive behaviour:

> “We see adolescent boys who are using abuse against their girlfriends but who have experienced a lot of childhood trauma and domestic violence as children and the CAMHS professionals tend to hold the boys’ trauma in mind over and above the trauma he’s causing.” (P22, domestic abuse)

Child and adult mental health services were the focus because this was often the service that users of abusive behaviour might already be in contact with, according to our participants, and they felt it was better for the work to start with a professional who was already in contact with the parent or young person. Support for people who use violence to change was seen as key to prevention of future domestic abuse both in adults but also in young victims who might go on to have their own harmful relationships.

## Discussion

This qualitative study explored professional perspectives of service co-ordination for families experiencing IPV and mental health problems. The systems mapping of interrelationships identified complex barriers and enablers to service co-ordination in the wider system through to the relationships between the frontline services themselves. A key gap was identified between domestic abuse services and statutory CYPMHS. Exploration of the boundaries of care suggested that children living in households with ongoing or intermittent IPV and children and parents with mental health or social care needs below or between service thresholds might be less able to access support. An overview of staff perspectives revealed differing views on addressing the effects of trauma, the co-ordination and sequencing of care, and service responsibility for parents who use abusive behaviour.

The key gap we identified between statutory CYPMHS services and voluntary sector domestic abuse services in part explains why children can be invisible in the wider domestic abuse support system. (28) This service gap has been highlighted across the voluntary sector service literature and reiterates recent calls for better integration between specialist IPV and mental health services (29) with joint commissioning as the central component. (17) Our lived experience group felt that there was a particular gap for teenage boys and that any link between domestic abuse services and CYPMHS would need to proactively address this.

This gap between CYPMHS and domestic abuse services appeared to be a specific manifestation of a deeper difference in perspectives between these services relating to how trauma is recognised and addressed in mental health services, (29) and how services can support children who are not considered to be in a position of safety. This is an ongoing debate around CYMPHS and their ability to provide therapeutic support for children with unstable home lives. An analysis of CYPMHS policy argues that a move towards individualised understandings of mental health excludes consideration of social conditions. (30) Earlier research with adult survivors of IPV had similar critiques of mental health services (31) and was echoed by our lived experience group who felt that CYPMHS can have a narrow perspective that does not relate mental health to the context a child might be living in. This professional separation between mental health and IPV is also reflected in child safeguarding policy: a recent review examined the representation of domestic abuse and found that mental health is rarely mentioned. (32)

This disciplinary and service separation between parental IPV and child mental health is also reflected in our finding that children (and parents) often fell between lower levels of mental health support and specialist mental health services. This has been widely reported as an issue across mental health services for children with more complex needs. For example, a recent evaluation of mental health support teams in schools (classed as a lower level of mental health support) found that children who were underserved by this service included children with special educational needs, those from ethnic minority backgrounds, and children with challenging family circumstances, including parental IPV. (33) Therapists in the study, like our participants, felt that the time-limited, low-intensity CBT on offer was not suited to these children, who also did not meet thresholds for more specialist support. (33) This suggests a wider systemic problem with service thresholds and the support offer affecting children living with social adversities.

### Strengths and limitations

Whilst this study incorporated the views of a range of health and domestic abuse professionals across three geographically varied areas in England, primary care was under-represented in our sample. Further work is needed to understand primary care practitioner perspectives; this is important given their central role in referring parents and children. However, resonance between our findings and national surveys (17,34), suggests the findings are more widely transferable beyond the three study sites. Research involving a wider range of health services and local authority services would enable further understanding of these perspectives, and in-depth work is required on the recognition of and support for IPV in CYPMHS.

### Implications for policy and practice

Our findings show that system-level intervention might be needed to improve service co-ordination, consistent with the wider health and domestic abuse service literature. Evaluation of Independent Domestic Violence Advisors (IDVA) in maternity services highlighted ongoing funding and local area health commissioning and management support were needed for service co-ordination. (35) Similarly, national reviews of services have highlighted the impact of commissioning decisions, (17,36) funding cycles (34,37) and funding restrictions, (38) and missing health service input in strategic meetings (34) on cross-sector service co-ordination. A review of health practitioner readiness to address IPV emphasised the need for supportive systems, (39) whilst the recent Lancet Psychiatry Commission on IPV highlighted trauma-informed support is only possible with system-wide changes. (29) Thus, attention needs to be paid to funding, commissioning and contracting processes, and the involvement of strategic groups, particularly smaller voluntary sector organisations and by and for services.

Our findings suggest there needs to be a strengthening of the relationship between domestic abuse services and CYPMHS including referrals and treatment pathways, in addition to cross-service advice and guidance. Currently, evidence about best practice is based on work with adults but suggests that there are pre-existing ways of working that could be expanded. Early work from the extension of the IRIS programme (domestic abuse advocates in general practice) has shown the success of extending the model to consider children (40) and this a possible model that could be developed in or with CYPMHS. A trial in the UK that trained domestic abuse advocates to support mental health had a positive impact on women’s mental health (41) suggesting that similarly this model could be extended to support children. Each of our study areas had an example of positive practice for children, these are described in supplement 6, Table 3.

Clinicians from mental health services consulting to the voluntary sector, suggested by our participants, might support the identification of families who fall through the service gap, in addition to strengthening the relationship between the sectors. Policy initiatives in the UK, such as Family Hubs and Early Help services, where health and social care services are already working together, provide pre-existing structures for strengthening the system.

## Conclusion

Improving service co-ordination for families experiencing IPV and mental health problems requires a systems approach to address the many barriers to accessing support parents and their children face. Improving the link between CYPMHS and domestic abuse services through ensuring relevant professionals from statutory and voluntary sector attend strategic groups, jointly contracting embedded workers across services and joint care pathways could go some way in addressing the main gaps in support.

## Data Availability

Data are not available due to confidentiality agreements.

## Acknowledgements

We wish to thank all the interview participants for giving their time to take part. We are grateful to Voices, our survivor advisory group and our three professional advisory groups for supporting this study and using their expertise to improve it.

## Supplementary Materials

### Supplement 1 – COREQ

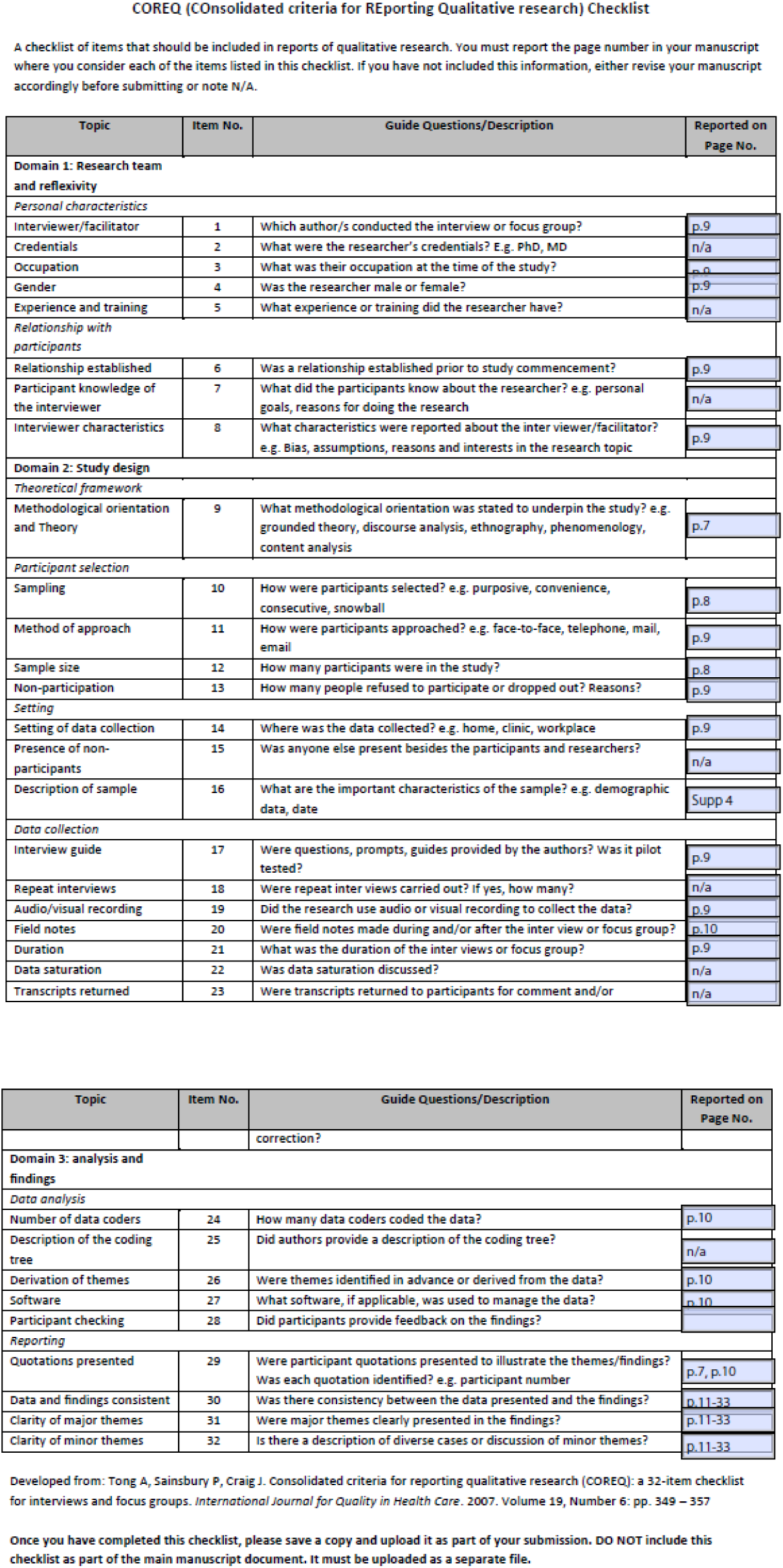

| Topic | Item No. | Guide Questions/Description | Reported on Page No. |
| --- | --- | --- | --- |
| **Domain 3: analysis and findings** |  |  |  |
| *Data analysis* |  |  |  |
| Number of data coders | 24 | How many data coders coded the data? | p.10 |
| Description of the coding tree | 25 | Did authors provide a description of the coding tree? | n/a |
| Derivation of themes | 26 | Were themes identified in advance or derived from the data? | p.10 |
| Software | 27 | What software, if applicable, was used to manage the data? | p.10 |
| Participant checking | 28 | Did participants provide feedback on the findings? |  |
| *Reporting* |  |  |  |
| Quotations presented | 29 | Were participant quotations presented to illustrate the themes/findings? Was each quotation identified? e.g. participant number | p.7, p.10 |
| Data and findings consistent | 30 | Was there consistency between the data presented and the findings? | p.11-33 |
| Clarity of major themes | 31 | Were major themes clearly presented in the findings? | p.11-33 |
| Clarity of minor themes | 32 | Is there a description of diverse cases or discussion of minor themes? | p.11-33 |

Developed from: Tong A, Sainsbury P, Craig J. Consolidated criteria for reporting qualitative research (COREQ): a 32-item checklist for interviews and focus groups. *International Journal for Quality in Health Care*. 2007. Volume 19, Number 6: pp. 349 – 357
Once you have completed this checklist, please save a copy and upload it as part of your submission. DO NOT include this checklist as part of the main manuscript document. It must be uploaded as a separate file.

### Supplement 2 – Changes to the protocol

We made the following changes due to recruitment difficulties:

- We moved away from a case study approach to a more general interview study, recruiting from three contrasting sites.
- We reduced our target sample size for primary care practitioners in each area from five to two and we reduced the interview schedule from 60 minutes to 30 minutes to encourage participation.
- We recruited from all three sites in parallel rather than one by one.
- We did not observe relevant team meetings as originally planned.

From the initial qualitative findings, it became clear that there were many moving parts to service coordination, and assessing these interconnected components and interrelationships called for a system complexity-aware analysis, rather than our original suggestion of applying candidacy framework and ecological systems theory. OA and CP embarked on this exercise by considering the gathered data through a systems lens. It is our view that this systems approach has greatly signified the missing connections and enhanced understandings.

### Supplement 3 – glossary and definitions

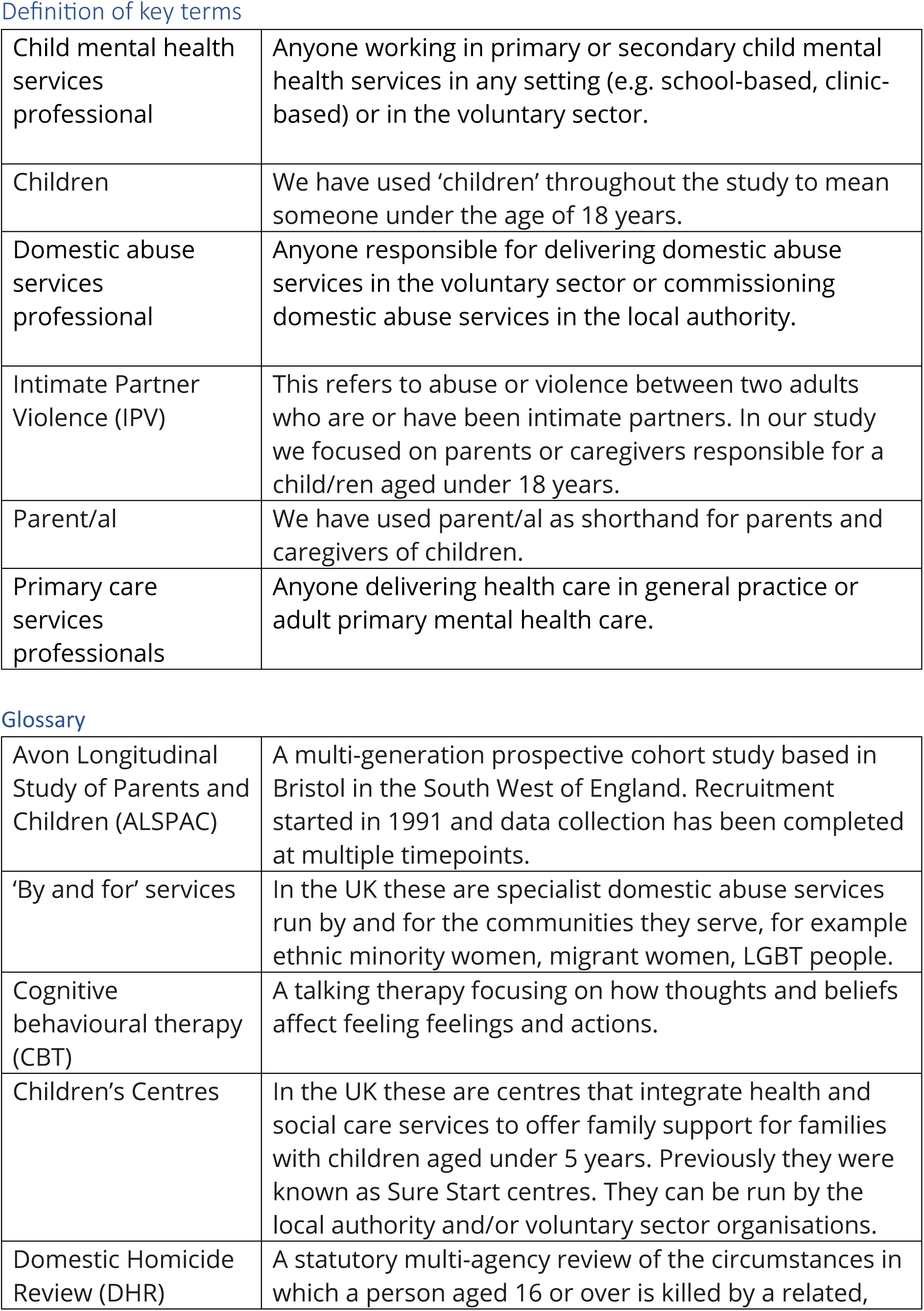
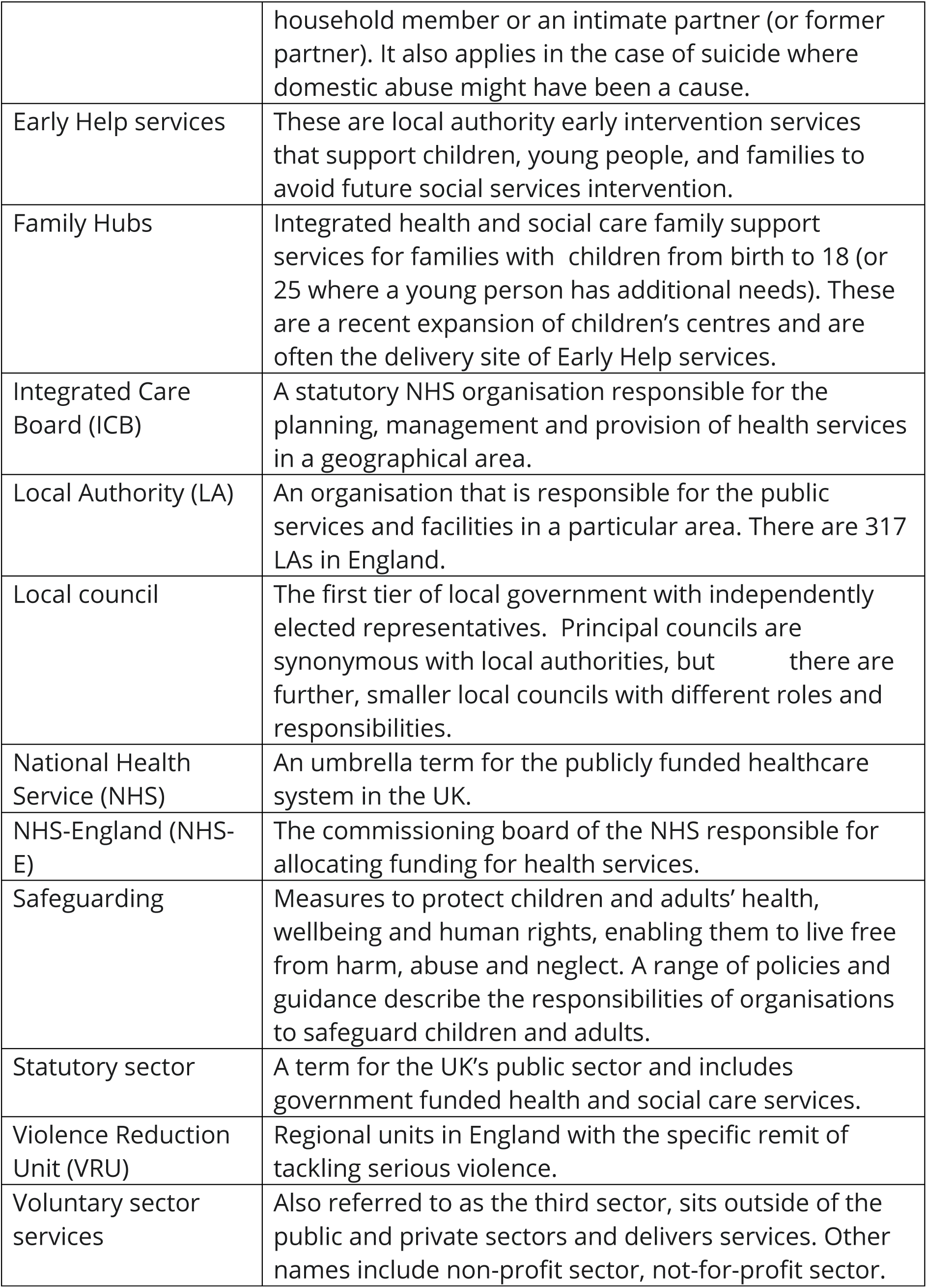

Definition of key terms
|  |  |
| --- | --- |
| Child mental health services professional | Anyone working in primary or secondary child mental health services in any setting (e.g. school-based, clinic-based) or in the voluntary sector. |
| Children | We have used 'children' throughout the study to mean someone under the age of 18 years. |
| Domestic abuse services professional | Anyone responsible for delivering domestic abuse services in the voluntary sector or commissioning domestic abuse services in the local authority. |
| Intimate Partner Violence (IPV) | This refers to abuse or violence between two adults who are or have been intimate partners. In our study we focused on parents or caregivers responsible for a child/ren aged under 18 years. |
| Parent/al | We have used parent/al as shorthand for parents and caregivers of children. |
| Primary care services professionals | Anyone delivering health care in general practice or adult primary mental health care. |
| Glossary |  |
| Avon Longitudinal Study of Parents and Children (ALSPAC) | A multi-generation prospective cohort study based in Bristol in the South West of England. Recruitment started in 1991 and data collection has been completed at multiple timepoints. |
| 'By and for' services | In the UK these are specialist domestic abuse services run by and for the communities they serve, for example ethnic minority women, migrant women, LGBT people. |
| Cognitive behavioural therapy (CBT) | A talking therapy focusing on how thoughts and beliefs affect feeling feelings and actions. |
| Children's Centres | In the UK these are centres that integrate health and social care services to offer family support for families with children aged under 5 years. Previously they were known as Sure Start centres. They can be run by the local authority and/or voluntary sector organisations. |
| Domestic Homicide Review (DHR) | A statutory multi-agency review of the circumstances in which a person aged 16 or over is killed by a related, |
|  | household member or an intimate partner (or former partner). It also applies in the case of suicide where domestic abuse might have been a cause. |
| Early Help services | These are local authority early intervention services that support children, young people, and families to avoid future social services intervention. |
| Family Hubs | Integrated health and social care family support services for families with children from birth to 18 (or 25 where a young person has additional needs). These are a recent expansion of children's centres and are often the delivery site of Early Help services. |
| Integrated Care Board (ICB) | A statutory NHS organisation responsible for the planning, management and provision of health services in a geographical area. |
| Local Authority (LA) | An organisation that is responsible for the public services and facilities in a particular area. There are 317 LAs in England. |
| Local council | The first tier of local government with independently elected representatives. Principal councils are synonymous with local authorities, but there are further, smaller local councils with different roles and responsibilities. |
| National Health Service (NHS) | An umbrella term for the publicly funded healthcare system in the UK. |
| NHS-England (NHS-E) | The commissioning board of the NHS responsible for allocating funding for health services. |
| Safeguarding | Measures to protect children and adults' health, wellbeing and human rights, enabling them to live free from harm, abuse and neglect. A range of policies and guidance describe the responsibilities of organisations to safeguard children and adults. |
| Statutory sector | A term for the UK's public sector and includes government funded health and social care services. |
| Violence Reduction Unit (VRU) | Regional units in England with the specific remit of tackling serious violence. |
| Voluntary sector services | Also referred to as the third sector, sits outside of the public and private sectors and delivers services. Other names include non-profit sector, not-for-profit sector. |

### Supplement 4 - Table 2: Sampling framework, recruitment targets, professional group definitions

Interview sampling framework and recruitment targets
| Professional group | Site 1<br>N=recruited<br>(n=target) | Site 2<br>N=recruited<br>(n=target) | Site 3<br>N=recruited<br>(n=target) | **Total** |
| --- | --- | --- | --- | --- |
| Child mental health | 5 (5) | 2 (5) | 5 (5) | 12 |
| Domestic abuse | 9 (5) | 5 (5) | 4 (5) | 18 |
| Primary care | 2 (5) | 2 (5) | 2 (2) | 6 |
| **Total** | 16 | 9 | 11 | 36 |

(NB: two of the interviews were group interviews with two participants each)

### Supplement 5 - Summary of IPV identification and training data

IPV identification
|  | IPV Identification. | Mental health identification |
| --- | --- | --- |
| Child mental health (CMH) professionals | Some CMH professionals reported routinely asking about parental IPV during assessments. Voluntary sector CMH professionals were more likely to ask about family violence in their assessments. Many CMH professionals reported not routinely asking. They would either pursue if they thought it was happening or ask indirect questions about trauma experiences and the past. CMH professionals reported that mothers are more likely to disclose than young people. | CMH professionals described asking parents directly or that parents might disclose about their own mental health. They might ask about parental mental health in the assessment or presume there were difficulties from the parent's presentation. |
| Domestic abuse (DA) professionals | All DA professionals reported asking about the details of parental IPV in their assessments. | All DA professionals reported asking adults about mental health in their assessments. They would ask parents about their children's mental health, either as part of the initial assessment or later on when they had started working together. Child-focused services would ask children directly about their own mental health. |
| Primary care (PC) professionals | PC professionals described routine asking about IPV only in specific circumstances, for example following IRIS training or in postnatal checks. Sometimes IPV would be flagged by health visitors. PC professionals felt that it takes time for adults to trust making a disclosure and they were sometimes anxious about asking directly. PC professionals with responsibility for safeguarding reported screening child hospital attendance data for patterns and following up on these. However they reported challenges with coding IPV in notes. | PC professionals described carrying out mental health assessments with parents. Nobody mentioned identifying children's mental health problems. |

IPV training
|  | What training have you had on IPV and/or mental health? | What training would you like? |
| --- | --- | --- |
| Child mental health (CMH) professionals | CMH professionals reported that they only received IPV training as part of safeguarding. The exception was professionals based in the voluntary sector who were more likely to have had extensive training. | CMH professionals wanted general IPV/domestic abuse training and awareness raising that was delivered by domestic abuse organisations and went beyond safeguarding. They were interested in learning about whole family approaches to IPV, including how to work with the perpetrator and how to support families who stayed together. They wanted to know how to have specific and supportive conversations with children, parents, and families about domestic abuse, in particular saying 'the right thing'. |
| Domestic abuse (DA) professionals | DA professionals were generally likely to have received mental health training, although the extent of this varied. Several reported not having any training specific to children. | DA professionals wanted up to date mental health training reflecting current best practice and that covered different family mental health presentations related to domestic abuse, as well as mental health crisis safety planning. In terms of child mental health, DA professionals wanted regular training on domestic abuse and the mental health impact on children as well as more specialised training on the more complex trauma presentations in children that might appear like Autism, delayed speech, hyperactivity. In addition, DA professionals wanted specialist training from other agencies, e.g. substance use, and ongoing support for not colluding with perpetrators. One organisation with expertise in mental health and domestic abuse wanted 'train the trainer' training to learn how to train others effectively. Some participants suggested it was important for commissioners to have sufficient time to do relevant training, particularly those without a practice background. DA professionals also recommended general DA training for school staff, specialist training for social workers, and encouragement for male GPs to attend domestic-abuse focused training. |
| Primary care (PC) professionals | All PC professionals reported receiving training on IPV as part of safeguarding, although it was treated as separate from mental health. The exceptions were IRIS trained professionals who felt they had in-depth training. | PC professionals wanted general IPV training for those outside of IRIS. They wanted specific child-focused IPV and mental health training related to safeguarding. They also wanted training on what other services do and where to refer for IPV. Practitioners felt that IPV should be integrated throughout all healthcare practitioner training. PC professionals also wanted broader training on trauma awareness and the impact on health. One professional felt that the issue was more around applying the training and suggested more case reviews as a way to address this. Another felt that GPs need more professional curiosity, rather than training. |

### Supplement 6 – Table 3: Examples of best practice

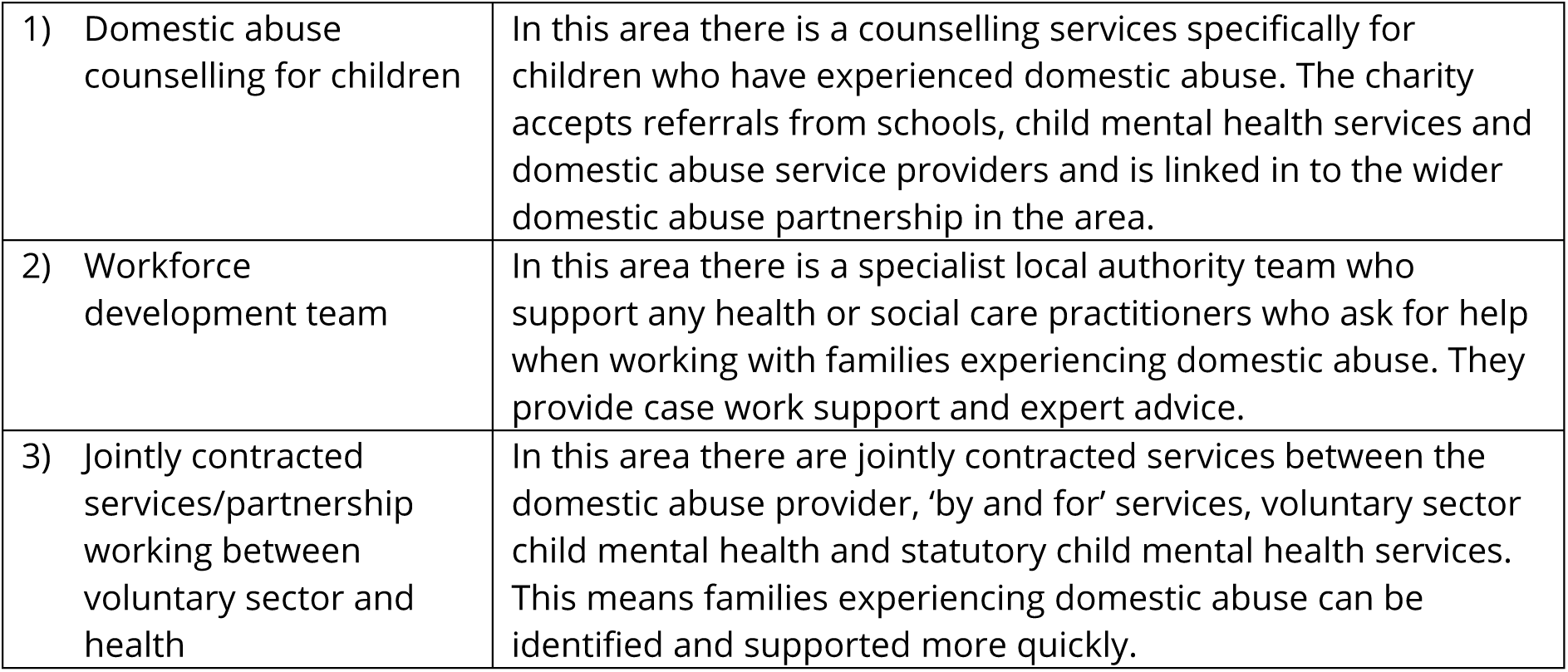

|  |  |
| --- | --- |
| 1) Domestic abuse counselling for children | In this area there is a counselling services specifically for children who have experienced domestic abuse. The charity accepts referrals from schools, child mental health services and domestic abuse service providers and is linked in to the wider domestic abuse partnership in the area. |
| 2) Workforce development team | In this area there is a specialist local authority team who support any health or social care practitioners who ask for help when working with families experiencing domestic abuse. They provide case work support and expert advice. |
| 3) Jointly contracted services/partnership working between voluntary sector and health | In this area there are jointly contracted services between the domestic abuse provider, 'by and for' services, voluntary sector child mental health and statutory child mental health services. This means families experiencing domestic abuse can be identified and supported more quickly. |

